# Associations of Genetic Variants in the Dopamine Transporter with Problematic Sexual Behavior and Reward Deficiency Syndrome

**DOI:** 10.1101/2024.09.21.24313023

**Authors:** Shui Jiang, Jerome C. Foo, Xiuying Hu, Leslie Roper, Esther Yang, Bradley Green, Randolph Arnau, Behavioral Addictions Studies and Insights Consortium, Rohit J. Lodhi, Rick Isenberg, David Wishart, Esther Fujiwara, Patrick J. Carnes, Katherine J. Aitchison

## Abstract

**Objectives:** Problematic sexual behavior (PSB) is defined by recurrent sexual behaviors that are difficult to control, causing social and functional impairments. PSB can co-occur with reward deficiency syndrome (RDS), but this relationship remains unclear. RDS has been associated with the 10/10 genotype of 3’ variable number tandem repeat (VNTR) in the dopamine transporter gene *SLC6A3,* which is implicated in the reward pathway. This study investigates the genetic relationship between PSB and RDS, testing their association with *SLC6A3* 3’ VNTR genotype.

**Methods:** PSB patients from addiction treatment facilities (n=454), and comparison participants (with non-clinical PSB=82; without PSB, n=888) were recruited. PSB was measured by the Sexual Addiction Screening Test-Revised (SAST-R) Core and RDS was measured using a composite variable from a custom test battery. DNA was collected from saliva and buccal swabs. Genotyping was performed using polymerase chain reaction (PCR), and regression analyses were conducted to investigate the association of *SLC6A3* 3’ VNTR genotype with PSB and RDS.

**Results:** The 10/10 genotype of *SLC6A3* 3’ VNTR was associated with RDS in a combined analysis of all groups, and with non-clinical PSB in the comparison participants. In patients, rare *SLC6A3* 3’ VNTR genotypes (3-, 6-, 8-, 11-repeat alleles) were associated with PSB. No genotype showed relationships to RDS in only PSB patients.

**Conclusions:** The association between the 10/10 genotype and RDS in the whole sample is consistent with previous findings associating this genotype with vulnerability to addictions. Consistent with our earlier report of PSB being related to RDS, this genotype was also associated with PSB in participants with PSB of a lesser severity. This is the first report of an association between the 10/10 genotype and PSB. In those with PSB of a greater severity, the power of the analysis was less, with a suggestive signal for an association between rare genotypes of the dopamine transporter and PSB.

## 1. INTRODUCTION

Problematic sexual behavior (PSB) is defined as difficult to control recurrent sexual behaviors that cause social and functional impairments (S. Jiang et al., 2023). PSB can be referred to as sex addiction (Carnes, 1991), or compulsive sexual behavior (Kuzma & Black, 2008). The scientific debate on the nosology and nature of PSB continues, but it is recognized that PSB often co-occurs with neuropsychiatric conditions such as reward deficiency syndrome (Blum et al., 2000; Comings & Blum, 2000), possibly suggesting common underlying biology.

Reward deficiency syndrome (RDS), often described as an “octopus of behavioral dysfunction,” is not a single mental health condition, but a collection of conditions including addictive, compulsive, and impulsive behaviors, and personality disorders, which share endophenotypes characterized by hypodopaminergia in the reward circuit (Blum et al., 2022; Blum et al., 2017; Gondre-Lewis et al., 2020). Although the relationship between RDS and PSB is not well characterized, there is emerging evidence that they may overlap (S. Jiang et al., 2023). Jiang et al. (2023) reported that compared to those without PSB, university students with PSB showed significantly higher rates of conditions with known alterations in the dopaminergic reward pathway which would fall under the umbrella of RDS (e.g., Attention Deficit/Hyperactivity Disorder [ADHD], obsessive-compulsive disorder [OCD], and personality disorders) (S. Jiang et al., 2023).

Addictive behaviors and symptoms are core features of RDS (Blum et al., 2024), and there are several genetic variants associated with vulnerability, maintenance, and relapse in addiction (Franklin et al., 2009; Ma et al., 2016; Reith et al., 2022). It is possible that at least part of the phenotypic overlap between PSB and RDS may be driven by genetic variants common to the two. For example, in one study, 81% of sexual addiction patients reported that at least one parent had an addictive disorder, with 36% indicating that a parent had a sexual addiction (Schneider & Schneider, 1996). One of the potential genetic variants for addictive disorder is the dopamine transporter gene *SLC6A3* (*solute carrier family 6 member 3*), located at chromosome 5p15.33. There is a 40-base pair variable number tandem repeat (VNTR) in the 3’ untranslated region of *SLC6A3* with 9- and 10- repeat alleles being the most common (Vandenbergh, et al., 1992a). Studies have shown that individuals with the *SLC6A3* 10/10 genotype express higher levels of dopamine transporter than 9-repeat allele carriers, and this is reflected in lower striatal dopamine among 10-repeat carriers (Dreher et al., 2009).

The dopamine transporter mediates the reuptake of dopamine from the synaptic cleft (Vandenbergh et al., 1992b), and the 10/10 genotype of the *SLC6A3* 3’ VNTR has been implicated in various conditions associated with an altered reward system, including ADHD and addictive disorders (Reith et al., 2022; Salatino-Oliveira et al., 2018; Volkow et al., 2011). For example, Cornish et al. (2005) reported that the 10/10 genotype was associated with higher ADHD symptom scores in boys (Cornish et al., 2005). A meta-analysis by Ma et al., (2016) showed that individuals carrying the 10/10 genotype were 17% less likely to quit smoking compared to those who was the 9-repeat alleles (Ma et al., 2016). Furthermore, a previous study indicated that the 10-repeat allele of the *SLC6A3* 3’ VNTR was associated with phenotypes directly related to sexual behavior. In a study of men, Guo et al. (2007) reported that individuals with at least one 10-repeat allele of the *SLC6A3* 3’ VNTR showed an 80%-100% increase in the number of sexual partners compared to those homozygous for the 9-repeat allele (Guo et al, 2008; Guo et al., 2007). While the 10/10 variant seems to be more consistently identified as the risk genotype, some studies probing gene-environment (G×E) interactions pointed to a greater sensitivity of 9-repeat carriers to environmental influences. For example, Erblich and colleagues (2004) reported that after stress, carriers of the 9-repeat allele showed a cigarette craving four times greater than non-carriers (Erblich et al., 2004). This contrasts with findings of potentially protective effects of the 10-repeat variant under adverse environmental influences (stress or childhood trauma) as seen in ADHD (Cornish et al., 2005), smoking (Ma et al., 2016), and number of sexual partners (Guo et al., 2008; Guo et al., 2007). However, these complex interactions have rarely been explored, and contradictory findings exist as well (Z. Jiang et al., 2023b).

Apart from common alleles such as 9- and 10-repeat, rare genetic variants, such as 3-, 5-, 7-, 8-, and 11-repeats are also observed (Vandenbergh et al., 1992a). Despite their low frequency and smaller contribution to population-wide heritability compared to common variants, these rare variants can still significantly increase individual risk of illness due to their pronounced impact on protein function or expression (Andreassen et al., 2023). A previous study including 471 children with or without ADHD found that *SLC6A3* rare genotypes (8/10, 7/10 and 10/11) were associated with ADHD (Šerý et al., 2015). However, research on these rare genotypes in candidate gene studies is limited, and these variants have often been excluded due to their low frequency and the resulting small sample sizes.

The aim of the present study was to investigate the relationship between PSB and RDS at the candidate gene level in PSB patients and in a non-clinical sample, with a focus on *SLC6A3* 3’VNTR genotype. We expected to extend the association between PSB and RDS that we had previously reported in the non-clinical comparison participants to a clinical sample. We hypothesized that the 10/10 genotype might be associated with PSB and/or with RDS.

## 1. METHODS

### 2.1. Participants

The clinical cohort comprised inpatients and outpatients receiving treatment for PSB at eight addiction treatment facilities in the United States (n=512). The recruitment of comparison participants (n=3530) was undertaken at a Canadian university as previously described (S. Jiang et al., 2023). Since the allele frequencies of the *SLC6A3* 3’ VNTR are observed to vary by ethnicity (Doucette-Stamm et al., 1995), genotypic and phenotypic analyses were restricted to individuals of European ancestry (n=1434, Supplementary Material, Figure 1).

### 2.2. Measures

We screened for PSB with the 20-item Sexual Addiction Screening Test-Revised (SAST-R) Core (Carnes et al, 2010). We measured four addictive dimensions of PSB: relationship disturbance caused by sexual behaviors (SAST-R Core items 6, 8, and 16, and we also included item 26 from the additional SAST-R subscales (Carnes et al, 2010), loss of control despite problems (SAST-R Core items 10, 12, 15, and 17), preoccupation about sex (SAST-R Core items 3, 18, 19 and 20), and affect disturbance (such as anxiety and depression related to sexual behaviors, SAST-R Core items 4, 5, 11, 13 and 14). Each question is responded to with a binary choice (yes/no=1/0), and the threshold for screening positive for PSB in the general population is 6 (Carnes et al, 2010). The PSB variable we used in all analyses (referred to as the “adjusted SAST-R Core”) was the scale total minus the first two questions, which inquire about a history of childhood sexual trauma and parental trouble with sexual behavior, as previously described (S. Jiang et al., 2023). These items were excluded due to their overlap with other variables in our analysis, such as “self-reported childhood trauma” and “family history of PSB.” This approach was taken to avoid redundancy and increase the specificity of the PSB regression, even though there are a part of SAST-R Core.

To derive our RDS measure, several relevant conditions were assessed, including nicotine dependence (Fagerström Test for Nicotine Dependence [FTND]), pathological gambling (DSM-5 Pathological Gambling Diagnostic Form [DPGDF]), personality disorders (8-item Standardized Assessment of Personality-Abbreviated Scale as a Self-Administered Screening Test [SA-SAPAS]), ADHD (Adult ADHD Self-Report Scale [ASRS] version 1.1), compulsive buying (Richmond Compulsive Buying Scale [RCBS]), and internet addiction (Internet Addiction Test [IAT]), as previously described (S. Jiang et al., 2023). To represent the overall RDS construct, a dummy coded binary variable was created (1=positive screening in any of the RDS components, 0=no). In addition, we collected self-reported information about demographic details, childhood trauma (self-reported childhood physical, emotional and sexual trauma, and family history of domestic violence, combined with item 1 from the SAST-R Core “sexually abused as a child”, 1=yes, 0=no, as previously described (S. Jiang et al., 2023)), past or current mental health diagnoses (1=yes, 0=no), number of past or current mental health diagnoses, and number of mental health condition(s) in the family history. A positive family history of PSB (1=yes, 0=no) was ascertained through item 2 of the SAST-R Core (“parental problems with sexual behavior”) and self-report data, as previously described (S. Jiang et al., 2023). Since OCD in particular is reported to be associated with PSB (S. Jiang et al., 2023) and RDS (Blum et al., 2000), we also used available data from MINI-International Neuropsychiatric Interview (MINI), in combination with self-reported past/current OCD diagnoses to estimate rates of OCD.

### 2.3. Procedures

DNA was extracted from saliva using Oragene OG-500 kits (DNAGenotek, Kanata, Canada) and/or from buccal swabs (Isohelix via D-Mark Biosciences, Toronto, Canada). Genotyping of the 40 bp *SLC6A3* 3’ VNTR was performed by a polymerase chain reaction (PCR). DNA was amplified with an initial denaturation step at 95°C for 2 minutes, followed by 35 cycles of denaturation at 95°C for 15 seconds, annealing at 64°C for 30 seconds, extension at 72°C for 1 minute, and a terminal extension step of 72°C for 5 minutes (modified from Vandenbergh et al., 1992) (Vandenbergh et al., 1992a). The primer sequences were: forward 5’-TGTGGTGTAGGGAACGGCCTGAG-3’, and reverse 5-CTTCCTGGAGGTCACGGCTCAAGG-3’ (Vandenbergh et al., 1992a). PCRs were resolved on a 2.25% agarose gel. The 9-repeat number was at 440 bp and the 10-repeat number was at 480 bp on the gel; genotypes other than 9/9, 9/10, or 10/10 are considered rare genotypes (Vandenbergh et al., 1992a). Since the allele frequencies of the *SLC6A3* 3’ VNTR vary by ethnicity (Doucette-Stamm et al., 1995), genotypic and phenotypic analyses were conducted in only those of European ancestry.

### 2.4. Statistical analysis

Data were analyzed using STATA (StataCorp, 2023). To examine the association of PSB, participants were categorized into three groups: (1) PSB patients (n = 405; patients who scored ≥11 on the unadjusted SAST-R Core); (2) Comparison participants with PSB (“non-clinical” PSB) (n=85) scoring ≥6 on the unadjusted SAST-R Core (Carnes et al., 2010), and (3) Comparison participants without PSB (n=439) scoring <6 on the unadjusted SAST-R Core and also screening negative on other relevant measurements (Carnes et al., 2010).

In the final analysis of PSB, a total of 929 participants (405 from the patient group and 524 from the comparison group) were included, after the removal of 59 individuals who were not of European ancestry or did not provide DNA. The SAST-R Core, developed based on a large clinical cohort (665 men and 274 women) of inpatients and outpatients in treatment for PSB, classifies individuals at a cut-point of 6, achieving a sensitivity of 0.817 and specificity of 0.778 (Carnes et al., 2010; Giordano et al., 2024). However, to enhance the specificity of the genetic association study, we selected a cut-off score of 11 on the SAST-R Core to focus on individuals with more pronounced PSB symptoms. This stricter threshold was implemented to ensure clearer signals for the genetic analysis by minimizing ambiguous cases, thereby prioritizing the identification of true positives over the risk of false negatives. Thus, we excluded 19 patients who scored <11 on the unadjusted SAST-R Core. In addition, to enhance the specificity of testing for associations with PSB, we excluded a total of 432 participants who screened negative for PSB, but positive for RDS or mental health conditions(s). This was done due to the consideration of potentially shared genetic etiologies between RDS, PSB, and mental health conditions. For the validation of RDS, all available data from patients (n=376) and comparison participants (n=668) were included.

#### 2.4.1. Descriptive statistics

To investigate if those with or without PSB have different clinical and demographic features, group differences were examined with Pearson’s χ^2^ test or a Mann-Whitney *U* test or a Kruskal-Wallis test, followed by Dunn’s test. To test if the distribution of *SLC6A3* genotype differed between different groups, Pearson’s χ^2^ was conducted. Tetrachoric (for binary variables) and Kendall’s τ (for ordinal variables) correlations were used to examine the correlation of clinical and demographic variables with severity of PSB.

#### 2.4.2. Regression analyses

To investigate the association of demographic and clinical variables with PSB and RDS, backward stepwise logistic regressions were performed. Regarding PSB, two regression models were performed: the first model included all comparison participants (with and without PSB), the second model included PSB patients and comparison participants without PSB. Initial independent variables in both models were: age, gender, *SLC6A3* genotype, childhood trauma, self-reported mental health condition(s) (a binary variable), number of mental health condition(s) in the family history (a continuous variable), and family history of PSB or of OCD. We also included an interaction term of childhood trauma and *SLC6A3* genotype to test for potential G×E interactions (Franke & Buitelaar, 2018; Z. Jiang et al., 2023; Lahey et al., 2011). To validate the association of the 10/10 genotype with RDS, one regression model was applied using the entire cohort. Initial independent variables were identical but additionally included “group” (comparison participants vs. patients). Predictors were retained in the models if the model passed the link test for correct specification and showed non-significant Hosmer-Lemeshow χ^2^, their inclusion did not introduce multicollinearity, and they remained significant in the final model. A *P*-value of <0.05 was considered significant.

### 2.5. Ethics

The study procedures were carried out in accordance with the Declaration of Helsinki. All participants provided written informed consent. The Quorum Review Institutional Review Board (Seattle, Washington; Protocol Number: 2016-001) and the University of Alberta Research Ethics Board (Protocol: Pro00066552) approved the study.

## 3. RESULTS

A total of 8.5% of comparison participants (82 out of 970) scored ≥ 6 on the unadjusted SAST-R Core, meeting the threshold for PSB in the general population (Carnes et al, 2010). Patients showed the highest scores in adjusted SAST-R Core and addictive dimensions of PSB (*P*<0.001, Table 1). Compared to comparison participants, patients were significantly older (*P*<0.001), had a higher proportion of male participants, and higher rates of RDS (*P*<0.001, Table 1). Furthermore, family history of mental health condition(s) and experiencing childhood trauma increased in frequency from comparison without PSB to comparison with PSB to PSB patients (*P*<0.001, Table 1).

**Table 1.** Demographic and Clinical Variables across Subgroups: Scores Indicate Median (Ranges) or Frequencies (Percentages)

| Variables |  | Comparison participants<br>without PSB<br>(n=266-448) | Comparison participants with<br>PSB<br>(n=55-89) | PSB Patients (n=418-434) |
| --- | --- | --- | --- | --- |
| Age <sup>a</sup> |  | 23 (18-55) <sup>b</sup> | 22 (18-66) <sup>b</sup> | 43 (19-80) <sup>c</sup> |
| Gender <sup>a</sup> | Women | 309 (69.8%) | 35 (42.2%) | 23 (6.3%) |
|  | Men | 134 (30.2%) | 50 (58.8%) | 408 (94.7%) |
| *SLC6A3*<br>genotype,<br>*P*=0.100 | 9/9 and 9/10 | 210 (46.9%) | 35 (39.3%) | 209 (48.2%) |
|  | 10/10 | 228 (50.9%) | 54 (60.7%) | 209 (48.2%) |
|  | Rare | 10 (2.2%) | 0 (0%) | 16 (3.6%) |
| Problematic Sexual Behavior -<br>adjusted SAST-R core <sup>a</sup> |  | 0 (0-5) <sup>b</sup> | 7 (4-18) <sup>c</sup> | 15 (10-18) <sup>d</sup> |
| *Relationship disturbance* <sup>a</sup> |  | 0 (0-2) <sup>b</sup> | 1 (0-4) <sup>c</sup> | 4 (0-4) <sup>d</sup> |
| *Loss of control* <sup>a</sup> |  | 0 (0-2) <sup>b</sup> | 2 (0-4) <sup>c</sup> | 4 (0-4) <sup>d</sup> |
| *Preoccupation* <sup>a</sup> |  | 0 (0-3) <sup>b</sup> | 1 (0-4) <sup>c</sup> | 2 (0-4) <sup>d</sup> |

| **Variables** | **Comparison participants**<br>**without PSB**<br>**(n=266-448)** | **Comparison participants with**<br>**PSB**<br>**(n=55-89)** | **PSB Patients (n=418-**<br>**434)** |
| --- | --- | --- | --- |
| *Affect disturbance* <sub>a</sub> | 0 (0-3) <sub>b</sub> | 3 (1-5) <sub>c</sub> | 4 (2-5) <sub>d</sub> |
| Reward deficiency syndrome <sub>a</sub> | 0 (0%) | 60 (67.4%) | 318(84.1%) |
| *FTND* <sub>a</sub> | 0 (0-1) <sub>b</sub> | 0 (0-7) <sub>c</sub> | 0 (0-12) <sub>c</sub> |
| *DPGDF (P=0.051)* | 0 (0-3) <sub>b</sub> | 0 (0-9) <sub>c</sub> | 0 (0-9) <sub>c</sub> |
| *SA-SAPAS* <sub>a</sub> | 1 (0-3) <sub>b</sub> | 1 (0-8) <sub>c</sub> | 2 (0-8) <sub>d</sub> |
| *ASRS* <sub>a</sub> | 1 (0-3) <sub>b</sub> | 3 (0-6) <sub>c</sub> | 3 (0-6) <sub>c</sub> |
| *RCBS* <sub>a</sub> | 10 (6-24) <sub>b</sub> | 11 (6-40) <sub>c</sub> | 11 (6-42) <sub>c</sub> |
| *IAT* <sub>a</sub> | 22 (0-30) <sub>b</sub> | 30 (0-62) <sub>c</sub> | 38 (0-97) <sub>d</sub> |
| Childhood trauma <sub>a</sub> | 67 (15.1%) | 33 (37.1%) | 290 (70.9%) |
| Self-reported mental health<br>condition(s) <sub>a</sub> | 0 (0%) | 24 (27.0%) | 277 (65.2%) |
| Number of self-reported mental<br>health condition(s) <sub>a</sub> | 0 <sub>b</sub> | 0 (0-12) <sub>c</sub> | 1 (0-9) <sub>d</sub> |
| Number of mental health<br>condition(s) in family history <sub>a</sub> | 3 (0-17) <sub>b</sub> | 4 (0-17) <sub>c</sub> | 6 (0-24) <sub>d</sub> |
| Family history of PSB <sub>a</sub> | 17 (3.8%) | 19 (21.6) | 273 (62.8%) |
| OCD <sub>a</sub> | 0 (0%) | 8 (9.0%) | 61 (14.1%) |

<sub>a</sub>: significant omnibus effect ( $P < 0.001$ );
<sub>b,c,d</sub>: different superscripts denote significant differences ( $P < 0.05$ ) between groups
Kruskal-Wallis $H$ test and Dunn's post-hoc tests for continuous variables; Pearson's or Fisher's $\chi^2$ test (cells $< 5$ ) for categorical variables; Abbreviations: *PSB*: problematic sexual behavior; *SAST-R*: Sexual Addiction Screening Test-Revised; *ASRS*: Adult ADHD Self-Report Scale version 1.1; *FTND*: Fagerström Test for Nicotine Dependence; *IAT*: Internet Addiction Test; *OCD*: obsessive-compulsive disorder; *RCBS*: Richmond Compulsive Buying Scale

The distributions of *SLC6A3* genotypes (9/9, 9/10, and 10/10) were in Hardy-Weinberg Equilibrium in both comparison participants and in patients (Supplementary Material, Table 1). Results showed that comparison participants with the 10/10 genotype had trend level elevated rates of PSB compared to 9-repeat carriers (*P*=0.050, Table 2). They also displayed significantly higher RDS total scores (*P*=0.0042), especially regarding ADHD symptoms (ASRS, *P*=0.021) and internet addiction (IAT, *P*=0.0073), followed by a trend level elevation in compulsive buying (RCBS, *P*=0.050).

**Table 2.** Distribution of Demographic and Clinical Variables by *Solute Carrier Family 6 Member 3 (SLC6A3)* Genotypic Group: Scores Indicate Median (Ranges) or Frequencies (Percentages)

| Variables | Comparison participants<br>(n= 55-89 with PSB; n=266-438<br>without PSB) |  | PSB patients (391-418) and comparison<br>participants without PSB (n=266-438) |  |  |
| --- | --- | --- | --- | --- | --- |
|  | 9/9 and 9/10 | 10/10 | 9/9 and 9/10 | 10/10 | Rare |
|  | genotypes) | genotype | genotypes | genotype | genotypes |
| PSB | 30 (12.2%)<sub>a</sub> | 52 (18.4%)<sub>a</sub> | 200 (48.1%) | 202 (46.1%) | 16 (61.5%) |
| Adjusted SAST-R core | 1 (0-14) | 1 (0-18) | 4.5 (0-18) | 3 (0-18) | 12.5 (0-18) |
| *Relationship disturbance* | 0 (0-3) | 0 (0-4) | 3 (0-4) | 3 (0-4) | 3 (0-4) |
| *Loss of control* | 0 (0-4) | 0 (0-4) | 2 (0-4) | 1 (0-4) | 3 (0-4) |
| *Preoccupation* | 0 (0-4) | 0 (0-4) | 1 (0-4) | 1 (0-4) | 1.5 (0-4) |
| *Affect disturbance* | 0 (0-4) | 0 (0-5) | 2 (0-5) | 2 (0-5) | 4 (0-5) |
| Reward deficiency syndrome | 21 (8.6%)<sub>a</sub> | 39 (13.8%)<sub>a</sub> | 155 (38.3%) | 152 (36.4%) | 11(45.8%) |
| *FTND* | 0 (0-4) | 0 (0-7) | 0 (0-12) | 0 (0-11) | 0 (0-4) |
| *DPGDF* | 0 (0-9) | 0 (0-9) | 0 (0-8) | 0 (0-9) | 0 (0-3) |
|  | 9/9 and 9/10 | 10/10 | 9/9 and 9/10 | 10/10 | Rare |
|  | genotypes) | genotype | genotypes | genotype | genotypes |
| *SA-SAPAS* | 1 (0-7) | 1 (0-8) | 1 (0-8) | 1 (0-8) | 1 (0-6) |
| *ASRS* | **1 (0-6)** <sub>b</sub> | **1 (0-6)** <sub>b</sub> | 2 (0-6) | 2 (0-6) | 1 (0-6) |
| *RCBS* | **10 (6-30)** <sub>b</sub> | **10 (6-40)** <sub>b</sub> | 10 (6-32) | 10 (6-32) | 10 (6-42) |
| *IAT* | **22 (0-55)** <sub>b</sub> | **24 (2-62)** <sub>b</sub> | 25 (0-89) | 26 (1-97) | 28 (4-88) |
| Childhood trauma | 45 (18.4%) | 55 (19.7%) | 169 (41.7%) | 176 (41.7%) | 12 (46.1%) |
| Self-reported mental health condition(s) | 7 (2.9%) <sub>a</sub> | 17 (6.0%) <sub>a</sub> | 131 (31.7%) | 134 (30.9%) | 12 (48.0%) |
| Number of self-reported mental health<br>condition(s) | 0 (0-10) <sub>a</sub> | 0 (0-12) <sub>a</sub> | 0 (0-6) | 0 (0-9) | 0 (0-9) |
| Number of mental health condition(s) in<br>family history | 3 (0-17) | 3 (0-17) | 4 (0-24) | 4 (0-19) | 4.5 (0-14) |
| Family history of PSB | 15 (6.1%) | 21 (7.5%) | 142 (33.9%) | 138 (31.6%) | 10 (38.5%) |
|  | 9/9 and 9/10<br>genotypes) | 10/10<br>genotype | 9/9 and 9/10<br>genotypes | 10/10<br>genotype | Rare<br>genotypes |
| OCD | 3 (1.2%) | 5 (1.8%) | 27 (6.5%)<sub>b</sub> | 29 (6.6%)<sub>b</sub> | 5 (19.2%)<sub>b</sub> |

<sup>a</sup>: $P < 0.1$ ;
<sub>b</sub>: $P < 0.05$
Mann-Whitney $U$ test for two-group comparisons of continuous variables; Kruskal-Wallis $H$ test and Dunn's post-hoc tests for three-group comparisons of continuous variables; Pearson's or Fisher's $\chi^2$ test (cells < 5) for categorical variables; Abbreviations: *PSB*: problematic sexual behavior; *SAST-R*: Sexual Addiction Screening Test-Revised; *ASRS*: Adult ADHD Self-Report Scale version 1.1; *FTND*: Fagerström Test for Nicotine Dependence; *IAT*: Internet Addiction Test; *OCD*: obsessive-compulsive disorder; *RCBS*: Richmond Compulsive Buying Scale

In comparison participants, the 10/10 genotype showed trend level correlations with RDS (τ=0.077, *P*=0.088), with self-reported mental health condition(s) (τ=0.081, *P*=0.073), as well as with PSB (τ=0.082, *P*=0.067; Supplementary Material, Figure 2). In patients, however, the 10/10 genotype was not significantly correlated with PSB or RDS. Our results higher rates of OCD in *SLC6A3* 3’ VNTR rare genotype carriers (*P*=0.044, Table 2). Additional analyses on correlations between genotype and SA-SAPAS items are shown in Supplementary Material, Table 2.

### Regression analyses

In both regression models for PSB, *SLC6A3* genotype, childhood trauma, family history of PSB, and gender were retained as independent variables (Model 1 in comparison participants: pseudo *R^2^*=0.12, n=509, Figure 1; Model 2 in PSB patients and comparison participants without PSB: pseudo *R^2^*=0.58, n=739, Figure 2). In Model 1, the 10/10 genotype of *SLC6A3* 3’ VNTR (OR: 1.97, 95% confidence interval (CI): 1.15-3.38, *P*=0.013), childhood trauma (OR: 2.31, 95% CI: 1.21-4.41, *P*=0.011), family history of PSB (OR: 4.49, 95% CI: 1.85-10.90, *P*<0.001) and male gender (OR: 4.54, 95% CI: 2.64-7.83, *P*<0.001) were associated with PSB. In Model 2, *SLC6A3* 3’ VNTR rare genotypes (OR: 5.69, 95% CI: 1.08-30.07, *P*=0.041), childhood trauma (OR: 9.78, 95% CI: 4.91-19.49, *P*<0.001), family history of PSB (OR:45.65, 95% CI: 15.56-133.90, *P*<0.001), and male gender (OR: 142.77, 95% CI: 52.11-391.15, *P*<0.001) were associated with PSB. The 10/10 genotype was not significantly associated with PSB in model 2 (*P*=0.72). The interaction term of childhood trauma and *SLC6A3* genotype was not significant in either comparison participants (*P*=0.36) or in patients (*P*=0.26).

**Figure 1.**
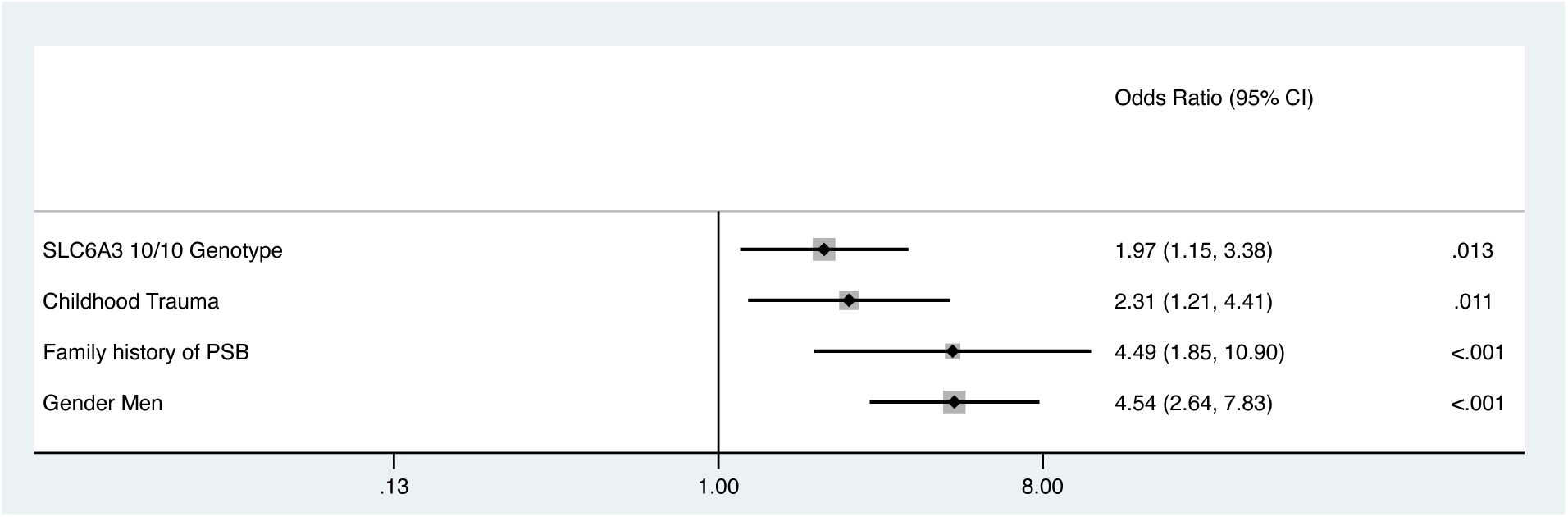
Forest Plots of Problematic Sexual Behavior (PSB) Logistic Regression Analysis in Comparison Participants. By logistic regression. The log likelihood of the model was -192.24 (n = 509, with 5 outliers removed, with a pseudo R^2^ = 0.12, *P*<0.001). The model passed the link test with a significant *P*(hat) (*P*=0.0080) and a nonsignificant *P*(hatsq) (*P*=0.66). The probability of the Hosmer-Lemeshow χ2 test was insignificant (*P*=0.75), suggesting good model fit. The mean VIF and the condition number were 1.06 and 3.38, respectively. The model accuracy, area under the ROC curve, sensitivity, specificity, and F score were 85.07%, 73.87%, 3.90%, 99.54%, and 0.073, respectively. The exact *P*-values were as follows: family history of PSB (*P*=9.19×10^-4^), gender (men: *P*=5.17×10^-8^). Power analysis demonstrated that the regression model had a power of over 0.90. *Abbreviation: PSB: problematic sexual behavior; SLC6A3: solute carrier family 6 member 3; VIF: variance inflation factor*.

**Figure 2.**
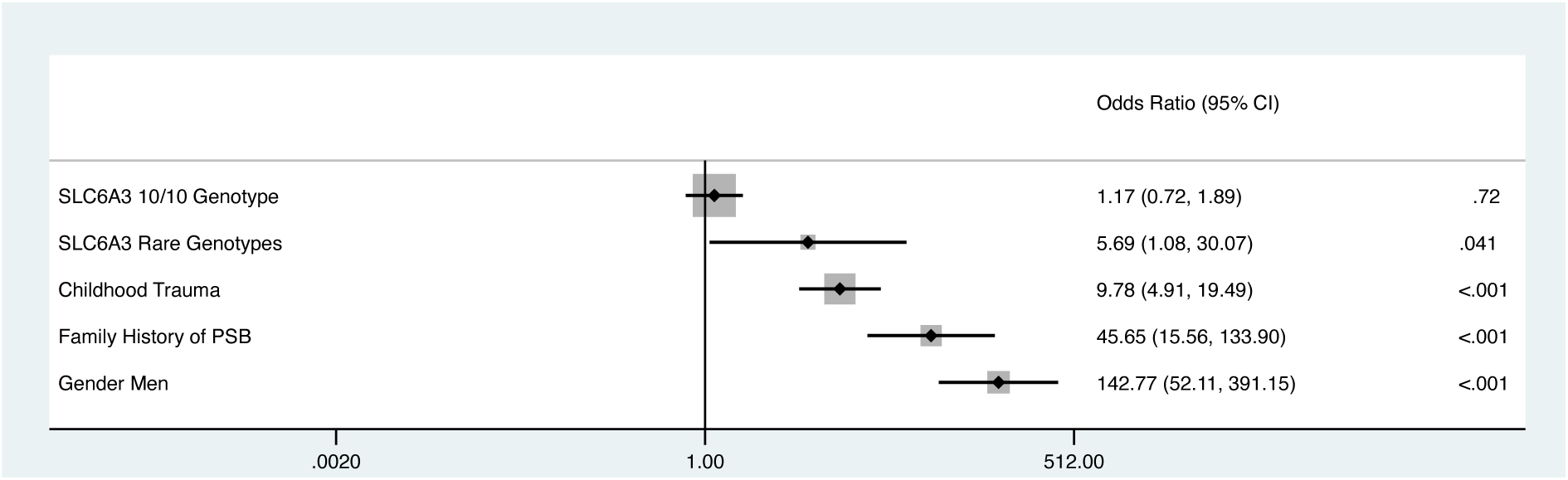
Forest Plots of Problematic Sexual Behavior (PSB) Logistic Regression Analysis in Patients and Comparison Participants Without PSB. By logistic regression. The log likelihood of the model was -195.53 (n=739, after removing outliers, n=105, with 102 having PSB, and with a pseudo R^2^=0.58, *P*<0.001). The model passed the link test with a significant *P*(hat) (*P*<0.001) and a nonsignificant *P*(hatsq) (*P*=0.56). The probability of the Hosmer-Lemeshow χ2 test was insignificant (*P*=0.98), suggesting a good model fit. The mean VIF and the condition number were 1.60 and 4.18, respectively. The model accuracy, area under the ROC curve, sensitivity, specificity, and F score were 85.39%, 94.03%, 70.63%, 95.64%, and 0.80, respectively. The exact *P*-values were as follows: childhood trauma (*P*=8.72×10^-11^), family history of PSB (*P*=3.43×10^-12^), and gender (men, *P*<2.00×10^-16^). Power analysis demonstrated that the regression model had a power of over 0.90. *Abbreviation: PSB: problematic sexual behavior; SLC6A3: solute carrier family 6 member 3; VIF: variance inflation factor*.

For RDS, the final model retained the interaction term of *SLC6A3* genotype and childhood trauma, self-reported mental health condition(s), *SLC6A3* genotype, childhood trauma, and group status on the entire cohort (pseudo *R^2^*=0.13, n=1044, Figure 3). Significant parameters included the 10/10 genotype of *SLC6A3* 3’ VNTR (OR:2.39, 95% CI:1.52-3.77, *P*<0.001), self-reported mental health condition(s) (OR:2.64, 95% CI: 1.96-3.56, *P*<0.001), childhood trauma (OR:2.81, 95% CI:1.72-4.60, *P*<0.001), and being a PSB patient (OR:4.10, 95% CI:2.96-5.68, *P*<0.001). The interaction term of the 10/10 genotype and childhood trauma was significantly associated with decreased odds of RDS (OR:0.32, 95% CI:0.17-0.60, *P*=0.011, the detailed interaction terms and predicted margins are provided in Supplementary Material, Table 3).

**Figure 3.**
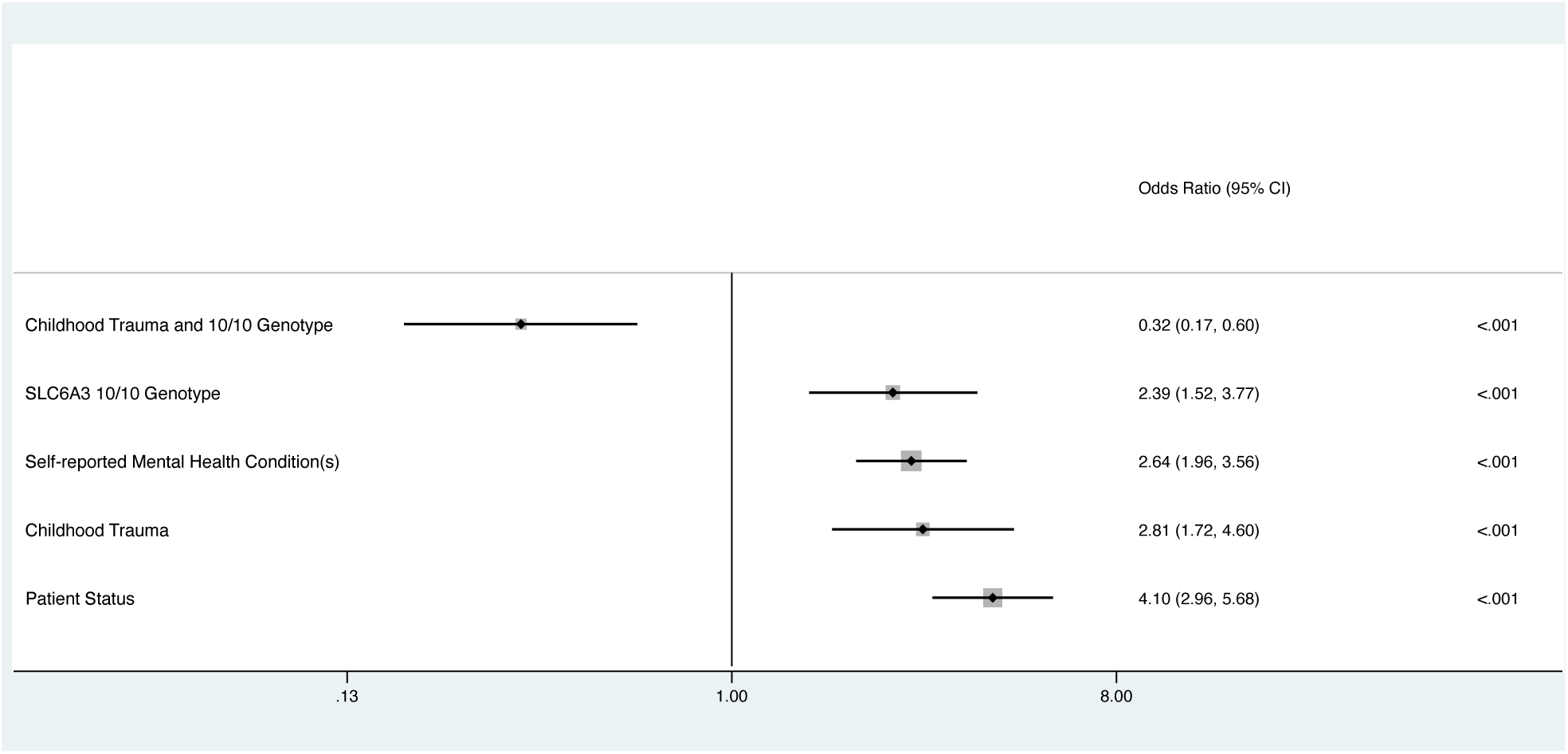
Forest Plot of Reward Deficiency Syndrome (RDS) Logistic Regression Analysis in Patients and Comparison Participants. By logistic regression. The log likelihood of the model was -624.29 (n=1044, after removed 282 outliers, with 91 of them being RDS positive, a pseudo *R^2^*=0.13, *P*<0.001). The model passed the link test with a significant *P*(hat) (*P*<0.001) and a nonsignificant *P* (hatsq) (*P*=0.81). The probability of the Hosmer-Lemeshow χ2 test was insignificant (*P*=0.99), suggesting a good model fit. The mean VIF and the condition number were 2.49 and 10.03, respectively. The model accuracy, area under the ROC curve, sensitivity, specificity, and F score were 68.29%, 72.00%, 67.46%, 69.16%, and 0.71, respectively. The exact *P*-values were as follows: the interaction term of 10/10 genotype and childhood trauma (*P*=3.68×10^-4^), the 10/10 genotype of *SLC6A3* 3’ VNTR (*P*=1.73×10^-4^), self-reported mental health condition(s) (*P*=1.46×10^-10^), childhood trauma (*P*=3.88×10^-5^) and being PSB patients (*P*<2.00×10^-16^). Power analysis demonstrated that the regression model had a power of over 0.90. *Abbreviation: PSB: problematic sexual behavior; SLC6A3: solute carrier family 6 member 3; VIF: variance inflation factor*

## 4. DISCUSSION AND CONCLUSION

In our study, the 10/10 genotype of the *SLC6A3* 3’ VNTR was associated with PSB in healthy young adults, and with RDS across the entire cohort, suggesting that this genotype may serve as a shared genetic risk factor for both PSB and RDS. This indicated that PSB could be a manifestation of RDS in this cohort of healthy young adults. Turner et al. (2022) indicated that PSB can develop as a side effect of dopaminergic agonists in some patients with Parkinson’s disease (Turner et al., 2022), aligning with our results, which aligns with our findings on the potential involvement of the dopaminergic pathway in both RDS and PSB. However, in the clinical cohort, it was rare genotypes, rather than the 10/10 genotype, that showed the association to PSB. This observation aligns with the understanding that both common and rare genetic variants can be associated with a psychiatric phenotype, with common genotypes typically exerting a smaller impact, while rare genotypes tend to have a larger impact and are associated with more severe symptoms (Andreassen et al., 2023).

Whether PSB should be regarded as a behavioral addiction or compulsive disorder remains an open question (Kraus et al., 2016; Walton & Bhullar, 2018). Of note, Koob and colleagues (2016) conceptualized addiction as a cycle with three stages, linked to disruptions in dopaminergic and serotoninergic pathways (Koob & Volkow, 2016). They further indicated that impulsivity is often seen in the early stages of addiction, and compulsivity evolves as the condition progresses (Koob & Volkow, 2016). Impulsive, reward-seeking tendencies of PSB (Kraus et al., 2016; Walton & Bhullar, 2018) could be more prominent in milder forms of PSB, such as in the comparison group, and appear to be associated with the relatively common 10/10 VNTR genotype. In contrast, the compulsive features may prevail in clinical PSB, accompanied by multiple mental health comorbidities (e.g., OCD), and then be associated with rare VNTR genotypes.

Through our convenience sampling we were not able to match comparison participants and PSB patients on age and other demographic variables. These group differences could have driven different manifestations of PSB, which may be associated with different genetic risk factors. Riemersma et, al. (2013) proposed a “new generation of sex addiction,” or “comtemporary sex addiction (Riemersma & Sytsma, 2013).” While both “classical sex addiction” and “contemporary sex addiction” could be assaociated with alterations in the reward pathway, Riemersma et, al. (2013) suggested that “contemporary sex addiction” is primarily linked to reward seeking, whereas “classical sex addiction,” is predominantly associated with trauma, emotional dysregulation, and insecure attachment, and may involve alterations in the prefrontal and limbic regions (Riemersma & Sytsma, 2013; Schmidt et al., 2017). In our study, comparison participants with PSB were significantly younger and a lower ratio of them reported childhood trauma than PSB patients. Hence, the relevance of the 10/10 genotype to RDS in the entire cohort might have been influenced by the larger proportion of younger participants who could have had such “contemporary” PSB. In contrast, rare genotypes of the *SLC6A3* 3’ VNTR, higher rates of childhood trauma and mental health comorbidities such as OCD could potentially be associated with more “classical” sex addiction in the clinical PSB group. Interestingly, our results suggest that in patients, this VNTR may have a potential link to OCD, with those who carried rare genotypes having higher rates of OCD. Some previous research has suggested a potential association between this VNTR and OCD (Billett et al., 1998; Hemmingsa et al., 2003); however, further studies are needed to confirm these associations. Our findings suggest that treatment stratification could be advantageous; individuals with RDS-related PSB and those with more severe clinical manifestations of PSB may benefit from tailored therapies that target different neurological underpinnings.

We also investigated specific G×E interactions (*SLC6A3* 3’ VNTR genotype and childhood trauma) in the context of hypodopaminergic phenotypes of PSB and RDS. Although such interactions have been explored in some relevant psychiatric conditions, particularly in substance use, the results have been inconsistent or inconclusive (Z. Jiang et al., 2023). A systematic review examining G×E between dopaminergic genes (*DRD2*, *DRD4*, and *SLC6A3*) and psychological, family, and socio-demographic factors in relation to substance abuse reported that five out of nine studies identified at least one significant interaction (Z. Jiang et al., 2023). In the present study, we observed that the 9-repeat allele could be associated with RDS when childhood trauma was present. It is possible that 9-repeat allele carriers may be more sensitive to environmental influences. For example, other research has found that children with conduct disorder and the 9/9 genotype showed stronger reductions in symptom severity in response to both negative or positive parenting practices, compared to 10-repeat allele carriers (Lahey et al., 2011).

This study had several limitations. The sociodemographic differences between the comparison participants and patients in these convenience samples are important to acknowledge; as mentioned above, age influences the phenotype and genetic susceptibility to many psychiatric symptoms, presumably including PSB and RDS (Grunblatt et al., 2019; Riemersma & Sytsma, 2013). Furthermore, the analysis was limited to those of European ancestry and included relatively few women. Also, the sample size for rare genotypes was limited, making it challenging to investigate the differential phenotypes between common and rare genotypes. Additionally, only one genetic variant in the *SLC6A3* gene and only one gene in the dopaminergic pathway were investigated in this study. It is possible that other genes, gene-gene interactions and G×E interactions may be associated with PSB and could have impacted the results. Thus, the findings should be interpreted with caution, and sample recruitment and matching should be optimized in future studies. These should also incorporate a broader range of common genetic variants or use a genome-wide approach, which would offer deeper insights into the genetic foundations of PSB.

Our results suggest that both common and rare variants of *SLC6A3* may be associated with PSB. The link of the 10/10 genotype to RDS suggests that PSB could be a manifestation of RDS in comparison participants, whereas rare *SLC6A3* variants might be associated with clinical PSB, together with comorbid mental health issues.

## Supporting information

Supplemental Digital Content

## Data Availability

All data produced in the present study are available upon reasonable request to the authors.

## Authors’ contribution

RI, RJL, PJC, and KJA contributed to securing funding. RI, RJL, BG, PJC, and KJA contributed to the study concept and design and ethics. RI and PJC oversaw the patient recruitment by the BASIC, supported in terms of data collection by BG and RA, and for sample collection by KJA, LR, and XH. The university participant recruitment was led by KJA, supported by LR and EY. XH and SJ contributed to the *SLC6A3* genotyping. SJ, JCF, DW, EF, and KJA contributed to data analysis, interpretation, and drafting and revising the paper.

## Ethics

All participants provided written informed consent. The Quorum Review Institutional Review Board (Seattle, Washington; Protocol Number: 2016-001) and the University of Alberta Research Ethics Board (Protocol: Pro00066552) approved the study.

## Acknowledgement

The authors would like to express their gratitude to Dawon Lee for assisting with the study concept, design, and ethics; to Beatriz Carvalho Henriques for writing assistance with the first version of the protocol for genotyping the *SCL6A3* 3’ VNTR; to May Yu for *SLC6A3* genotyping; to Pennie Carnes and Keanna Wallace for sample collection at the university; and to Dr. Diego Lapetina for laboratory management support and preliminary data analysis.

