## Supplemental Digital Content for "Associations of Genetic Variants in the Dopamine Transporter with Problematic Sexual Behavior and Reward Deficiency Syndrome"

**Short title:** *SLC6A3* Variants and Problematic Sexual Behavior

**Authors:**

Shui Jiang,, PhD^1^

Jerome C. Foo,, PhD^1,2,3^

Xiuying Hu,, BM, MSc^4^

Leslie Roper,, MSc, MC^1^

Esther Yang,, MSc^1^

Bradley Green,, PhD^5^

Randolph Arnau,, PhD^6^

Behavioral Addictions Studies and Insights Consortium*,

Rohit J. Lodhi,, PhD, FRANZCP, FRCPsych^1,7^

Rick Isenberg,, MD^8^

David Wishart,, PhD^9^

Esther Fujiwara,, PhD^1^

Patrick J. Carnes,, PhD^10^,

Katherine J. Aitchison,, BM BCh, PhD, FRCPsych^1,3,4,11^

**Affiliations:**

^1^Department of Psychiatry, Faculty of Medicine and Dentistry, College of Health Sciences, University of Alberta, Edmonton, AB, Canada

^2^Department of Genetic Epidemiology in Psychiatry, Central Institute of Mental Health, Mannheim, Germany

^3^Neuroscience and Mental Health Institute, University of Alberta, Edmonton, AB, Canada

^4^Department of Medical Genetics, Faculty of Medicine and Dentistry, College of Health Sciences, University of Alberta, Edmonton, AB, Canada

^5^Department of Psychology and Counseling, University of Texas at Tyler, Tyler, TX, USA

^6^School of Psychology, University of Southern Mississippi, Hattiesburg, MS, USA

^7^Department of Psychiatry, Schulich School of Medicine and Dentistry, Western University, London, ON, Canada

^8^Psychological Counselling Services, Scottsdale, AZ, USA

^9^Department of Biological Sciences, Faculty of Science, College of Natural and Applied Sciences, University of Alberta, Edmonton, AB, Canada

^10^Gentle Path at the Meadows, Wickenburg, AZ, USA

^11^The Psychiatry Section, Division of Clinical Sciences, Northern Ontario School of Medicine, Thunder Bay, ON, Canada

**Correspondence to:**

Katherine Aitchison, BM BCh, PhD, FRCPsych

Tel number: 780-492-4018

5-020, Katz Group Centre for Pharmacy and Health Research, Edmonton, AB, Canada

***Consortium authors in supplementary material**

**Funding Sources:**

The work herein was supported by an Alberta Centennial Addiction and Mental Health Research Chair and transitional funding (to KJA), Canada Foundation for Innovation (CFI), John R. Evans Leaders Fund (JELF) grant (32147 - Pharmacogenetic translational biomarker discovery), Alberta Innovation and Advanced Education Small Equipment Grants Program (to KJA), and a research grant and philanthropic support from the American Foundation for Addiction Research (to KJA). A Fulbright-Canada-Palix Foundation grant (to PJC) assisted with his contributions to study design, project management, and collaborative working. SJ was supported by an Alberta Innovates Postdoctoral Recruitment Fellowship.

**Authors’ contribution:**

RI, RJL, PJC, and KJA contributed to securing funding. RI, RJL, BG, PJC, and KJA contributed to the study concept and design and ethics. RI and PJC oversaw the patient recruitment by the BASIC, supported in terms of data collection by BG and RA, and for sample collection by KJA, LR, and XH. The university participant recruitment was led by KJA, supported by LR and EY. XH and SJ contributed to the *SLC6A3* genotyping.  SJ, JCF, DW, EF, and KJA contributed to data analysis, interpretation, and drafting and revising the paper.

**Conflicts of interest:**

While PJC was previously a Board member of the American Foundation for Addiction Research, he is no longer a member. Moreover, neither the Foundation nor any other of the funders played any role in study design, or in data analysis or interpretation thereof. All other authors declare no conflicts of interest.

**Ethics:**

All participants provided written informed consent. The Quorum Review Institutional Review Board (Seattle, Washington; Protocol Number: 2016-001) and the University of Alberta Research Ethics Board (Protocol: Pro00066552) approved the study.

**Abstract word count:** 244

**Manuscript word count (not including references, tables, or figure legends):** 3847

**Number of references:** 40 **Figure 1. Study Recruitment and Analysis Flow Chart for Comparison Participants and Patients**

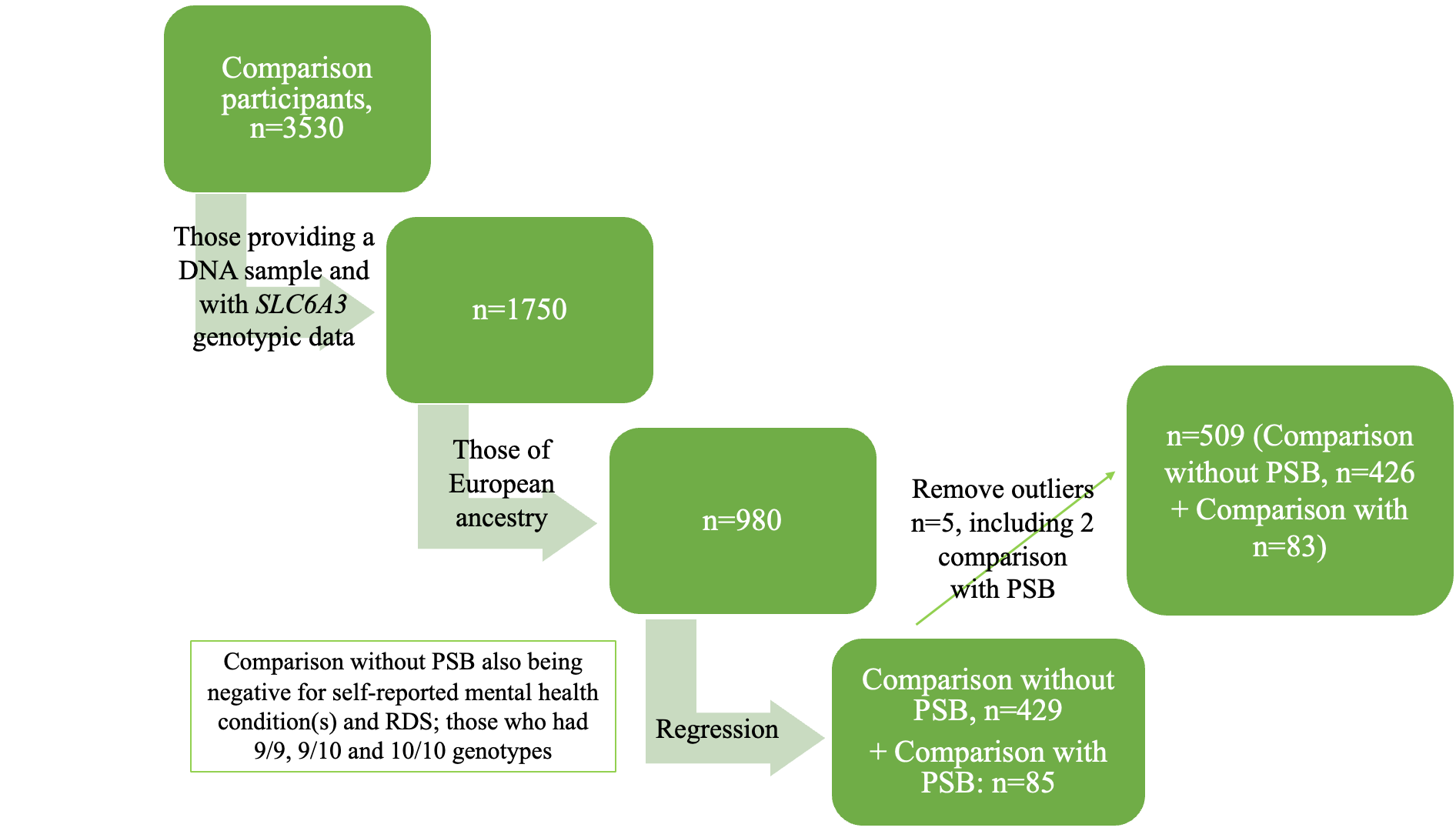

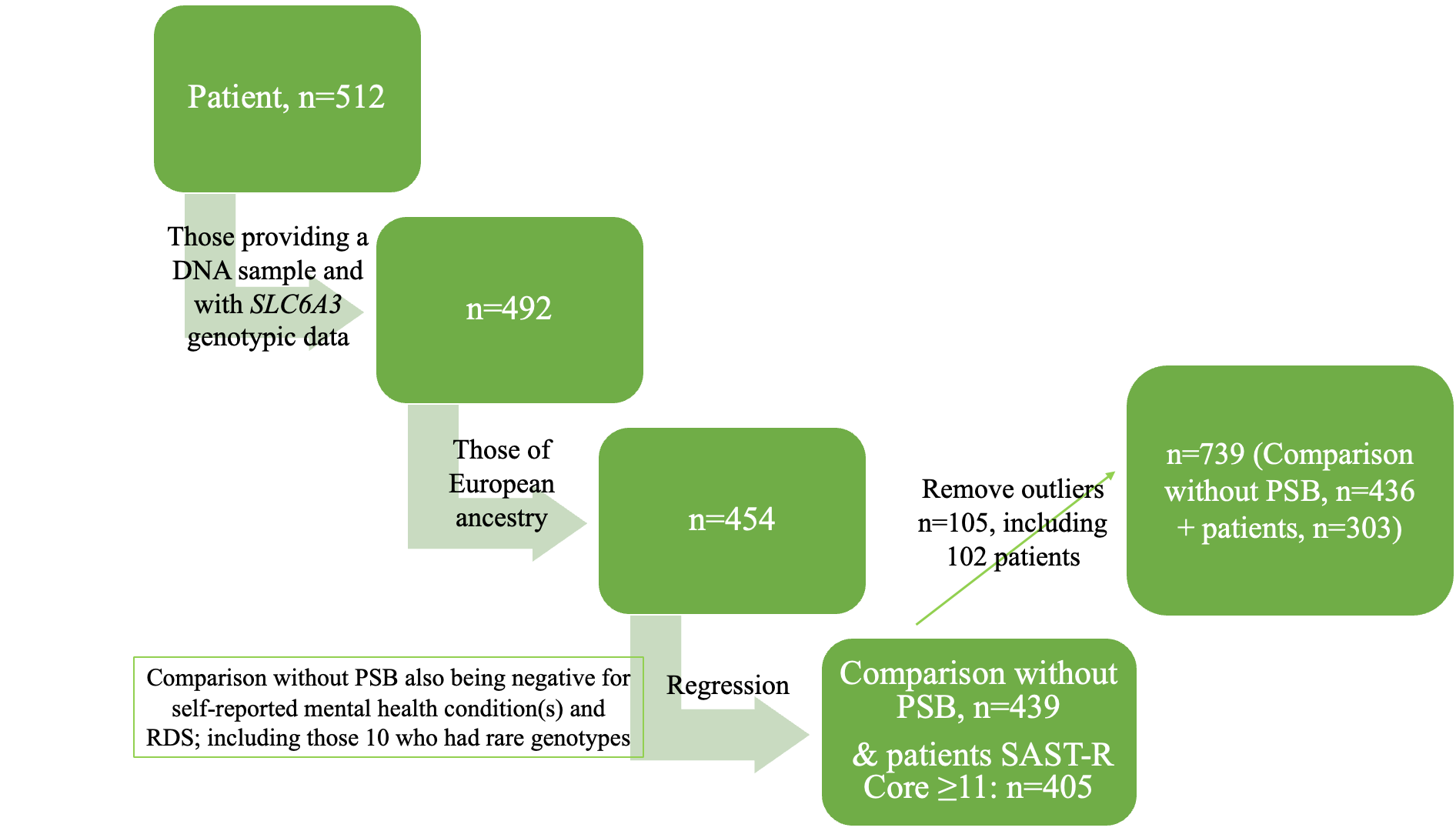

A. Logistic regression (n=509: 426 comparison without PSB and 83 with PSB; outliers: n=5, comparison participants, with 2 comparison with PSB) were conducted in comparison participants.

B. Logistic regression (n=739, 436 comparison without PSB and 303 patients; outliers: n=105, 3 comparison participants without PSB and 102 patients) were conducted in comparison without PSB and patients. *Abbreviation: PSB: problematic sexual behavior; SLC6A3: solute carrier family 6 member 3; SAST-R: Sexual Addiction Screening Test-Revised; RDS: reward deficiency syndrome.*

**Figure 2. Correlation Matrix for Problematic Sexual Behavior (PSB) with Demographic and Clinical Variables**

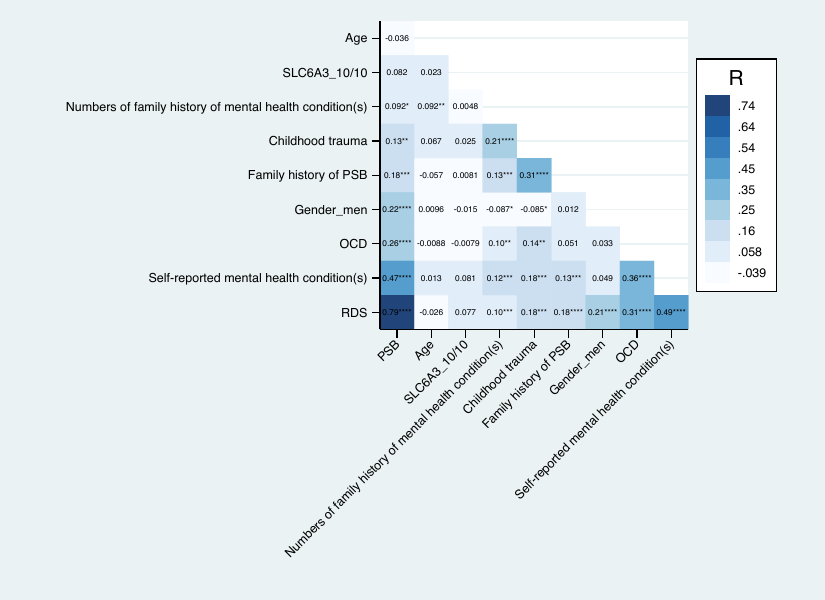

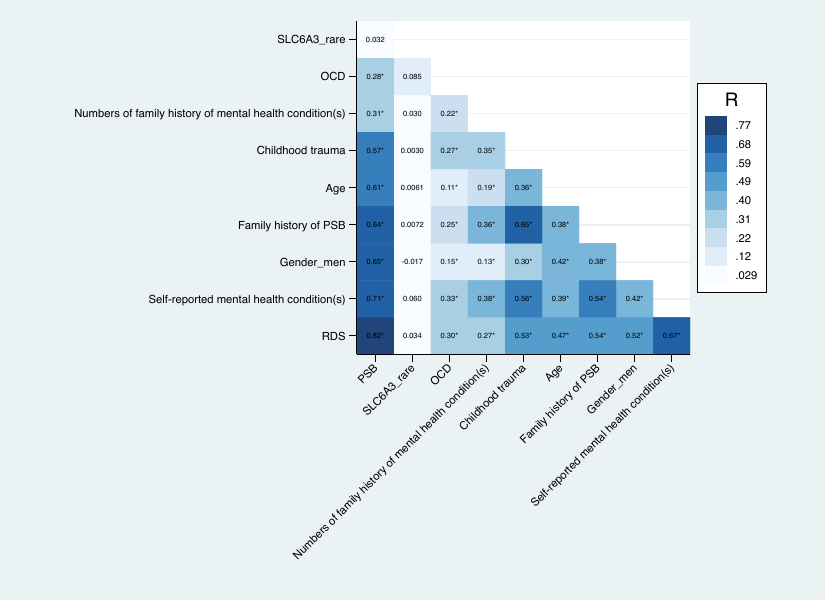

A. Correlation matrix for PSB in comparison participants by Kendall-τ correlation (n=491). *: *P*<0.05; **: *P*<0.01; ***: *P*<0.005; ****: *P*<0.001. B. Correlation matrix for PSB in patients vs. comparison without PSB by Kendall-τ correlation (n=769, including 420 comparison without PSB and 349 patients). *: significant omnibus effect (*P*<0.001).

*Abbreviation: PSB: problematic sexual behavior; SLC6A3: solute carrier family 6 member 3, SLC6A3 in comparison participants (9/9, 9/10 vs 10/10 genotype), SLC6A3 in patients (common vs rare genotypes); OCD: obsessive-compulsive disorder, combined MINI and self-report data; RDS: reward deficiency syndrome.*

**Table 1. Distribution of *Solute Carrier Family 6 Member 3 (SLC6A3)* Genotype in Comparison (A) and Patient (B) Participants with Hardy-Weinberg equilibrium (HWE) Test on the Data for the 9/9, 9/10, and 10/10 Genotypes**

| **Genotype** | | **Genotype frequency** | **Allele** | **Allele frequency** | **Expected genotype frequency** |
| --- | --- | --- | --- | --- | --- |
| Common | 9/9 | 64 (6.5%) | 9 | 0.26 | 65 |
|  | 9/10 | 374 (38.2%) |  |  | 372 |
|  |  |  | 10 | 0.74 |  |
|  | 10/10 | 528 (53.9%) |  |  | 529 |
| Rare (n=14, 1.4%) | 6/10 | 1 | *P*=0.98 _a_ | | |
|  | 7/10 | 1 |  |  |  |
|  | 7/9 | 1 |  |  |  |
|  | 8/10 | 1 |  |  |  |
|  | 9/11 | 4 |  |  |  |
|  | 10/11 | 5 |  |  |  |
| Total | | 980 |  |  |  |

| **Genotype** | | **Genotype frequency** | **Allele** | **Allele frequency** | **Expected genotype frequency** |
| --- | --- | --- | --- | --- | --- |
| Common | 9/9 | 28 (6.2%) | 9 | 0.28 | 34 |
|  | 9/10 | 188 (41.5%) |  |  | 176 |
|  |  |  | 10 | 0.72 |  |
|  | 10/10 | 221 (48.8%) |  |  | 227 |
| Rare (n=16, 3.5%) | 3/3 | 1 | *P*=0.36 _a_ | | |
|  | 6/9 | 1 |  |  |  |
|  | 6/10 | 2 |  |  |  |
|  | 8/9 | 3 |  |  |  |
|  | 8/10 | 3 |  |  |  |
|  | 9/11 | 1 |  |  |  |
|  | 10/11 | 5 |  |  |  |
| Total | | 453 |  |  |  |

_a_ Hardy-Weinberg equilibrium (HWE**)** Likelihood-ratio χ2 (2 alleles, 9 and 10)

A. In comparison participants (n=980). B. In patient (n=453*.* The 3 allele: 200 base pair (bp);

the 6 allele: 320 bp; the 8 allele: 400 bp; the 9 allele: 440 bp; the 10 allele: 480 bp; the 11 allele: 520 bp. *Abbreviation: SLC6A3: solute carrier family 6 member 3*

**Table 2. Correlation Matrix of the 8-item Standardized Assessment of Personality-Abbreviated Scale as a Self-Administered Test (SA-SAPAS) and Adjusted SAST-R Core Items by *Solute Carrier Family 6 Member 3 (SLC6A3)* Genotype**

| SA-SAPAS item | ρ, *P*-value |
| --- | --- |
| Having trouble making and keeping friends (item 1) | 0.091, ***P*=0.045** _a_ |
| Being a loner (item 2) | 0.15, ***P*<0.001** _a_ |

| SA-SAPAS item | ρ, *P*-value |
| --- | --- |
| Being a perfectionist (item 8) | -0.21, ***P*=0.034** _a_ |

| SAST-R Core addictive dimensions | ρ, *P*-value |
| --- | --- |
| Loss of control | 0.17, *P*=0.094 _a_ |
| Relationship disturbance | 0.20, ***P*=0.037** _a_ |

_a_ Tetrachoric correlation

A, The correlation of each SA-SAPAS item (0=no, 1=yes) and the 10/10 genotype, n=1401. B, The correlation of SA-SAPAS items and rare genotypes (common vs. rare genotypes), n=1430. C, The correlation of adjusted SAST-R Core items (0=no, 1=yes) and rare genotypes, n=1414

No significant correlations were found between *SLC6A3* genotype and other individual SA-SAPAS items (item 3: “do you trust other people;” item 4: “are you normally an impulsive sort of person;” item 6: “are you normally a worrier” and item 7: “do you depend on others a lot”). No significant correlations were found between *SLC6A3* genotype other PSB addictive dimensions (preoccupation, affect disturbance, and loss of control). *Abbreviation: SA-SAPAS: Standardized Assessment of Personality-Abbreviated Scale as a Self-Administered Test; SLC6A3: solute carrier family 6 member 3, SLC6A3 in comparison participants (9/9, 9/10 vs 10/10 genotype), SLC6A3 in patients (common vs rare genotypes); SAST-R: Sexual Addiction Screening Test-Revised.*

**Table 3.** **Predicted Margins of the Interaction Terms of *SLC6A3* Genotype and Childhood Trauma**

| **G×E interaction**  **(Childhood trauma ×**  ***SLC6A3* 3’ VNTR Genotype)** | **Predictive Margins (95% CI)** |
| --- | --- |
| No × 9/9 and 9/10 _a_ | 0.41 (0.33-0.48) |
| No × 10/10 _a_ | 0.59 (0.54-0.63) |
| Yes × 9/9 and 9/10 _a_ | 0.62 (0.55-0.69) |
| Yes ×10/10 _a_ | 0.57 (0.51-0.62) |

_a_: *P*<0.001

*Abbreviation: G×E :gene-environment interaction; SLC6A3: solute carrier family 6 member 3.*

**Detailed List of Family History of Mental Health Condition(s), and Self-reported Mental Health Condition(s)**

Family history of mental health condition(s): includes family history of intellectual disability, autism spectrum disorder, Attention-Deficit/Hyperactivity Disorder (ADHD), learning disability, Tourette Syndrome, schizophrenia, other psychotic disorder, depression, bipolar, anxiety, obsessive-compulsive disorder, post-traumatic stress disorder, dissociative identity disorder, anorexia, bulimia, sleep apnea, other sleep disorder, conduct disorder, alcoholism, drug addiction, nicotine addiction, gambling addiction, internet addiction, Alzheimer, other dementia, personality disorder, Parkinson’s and Huntington’s disease.

Self-reported mental health condition(s): includes the combination of previous and current diagnoses history of major depressive disorder, bipolar, anxiety, panic attack, post-traumatic stress disorder, eating disorder, psychosis, autism spectrum disorder, dissociative identity disorder, conduct disorder, body dysmorphic disorder, hoarding, trichotillomania, excoriation, and other psychotic disorder, intellectual disability, dementia, and cognitive impairment.

**The Behavioral Addictions Studies and Insights Consortium (BASIC)**

Kate Balestrieri, PsyD

Chris Chandler, MA, LMHC

Lauren Dummit, LMFT

Marcus Earle, PhD, LMFT

Greg Futral, PhD

Michelle Gaugh, MA

Piper Grant, PsyD, MPH

Alex Katehakis, PhD, MA, MFT

Barbara Levinson, PhD, RN, LMFT, LSOTP, CSAT Supervisor, CST Diplomate

Andrew Meadows, BS

Dan Morris, LCSW

Isabel Nino-de-Guzman, PhD

Helena Vissing, PsyD

Randolph Arnau, PhD*

Bradley Green, PhD*

Rick Isenberg, MD*

Patrick J. Carnes, PhD*

Katherine J. Aitchison, BM BCh, PhD, FRCPsych*

**lead investigator*

Affiliations

Triune Therapy Group, Los Angeles, CA, USA (KB, LD, HV)

Christian Health Group, La Jolla, CA, USA (CC)

Psychological Counseling Services, Scottsdale, AZ, USA (ME, RI)

Pine Grove Behavioral Health & Addiction Services, Hattiesburg, MS, USA (GF)

Center for Healthy Sex, Los Angeles, CA, USA (PG, AK)

Kavod Center, Rochester, NY, USA (MG, AM, DM)

Center for Healthy Sexuality, Houston, TX, USA (BL)

Gentle Path at the Meadows, Wickenburg, AZ, USA (ING, PJC)

Department of Psychology and Counseling, University of Texas at Tyler, Tyler, TX, USA (BG)

School of Psychology, University of Southern Mississippi, Hattiesburg, MS, USA (RA)

Department of Psychiatry, College of Health Sciences, University of Alberta, Edmonton, AB, Canada (KJA)

Department of Medical Genetics, College of Health Sciences, University of Alberta, Edmonton, AB, Canada (KJA)

Neuroscience and Mental Health Institute, University of Alberta, Edmonton, AB, Canada (KJA)

Women and Children’s Health Research Institute, University of Alberta, Edmonton, AB, Canada (KJA)

Psychiatry Section, Division of Clinical Sciences, Northern Ontario School of Medicine, Thunder Bay, ON, Canada (KJA)
